# Development and validation of Retrieval Augmented Generation (RAG) and GraphRAG for complex clinical cases

**DOI:** 10.1101/2025.11.25.25341010

**Authors:** M. Berkan Sesen, Joshua Au Yeung, Elham Asgari

## Abstract

**Objective:** Chronic Kidney Disease (CKD) is a progressive condition requiring evidence-based management, but adherence to complex guidelines remains challenging. Large Language Models (LLMs) could support clinical decision-making, yet their unreliability limits direct use. This study aimed to evaluate whether Retrieval-Augmented Generation (RAG), particularly a knowledge graph-enhanced pipeline (GraphRAG), improves guideline-based clinical decision support (CDS) in CKD management.

**Methods and Analysis:** We compared three approaches: a baseline LLM (GPT-4o), a vector-indexed RAG pipeline, and a GraphRAG pipeline. Each model answered nine clinically relevant questions for a synthetic cohort of 70 CKD patients. Outputs were assessed for clinical correctness, patient-specificity, and clarity, using both clinician-led evaluations and an LLM-as-Judge framework.

**Results:** RAG-based methods outperformed the baseline LLM in clinical correctness and guideline adherence. GraphRAG achieved the highest patient-specificity by leveraging multi-hop relationships across a knowledge graph derived from NICE CKD guidelines, particularly for tasks involving thresholds, algorithmic decisions, or open-ended management. However, GraphRAG scored lower in clarity, as its graph walks often returned long guideline excerpts that obscured key recommendations. All RAG systems were limited by the scope of the indexed guideline and performed poorly when essential information was missing.

**Conclusions:** RAG and GraphRAG provide a scalable, auditable foundation for guideline-aligned CDS in CKD, with GraphRAG showing particular strengths in tailoring advice to patient data. Nonetheless, trade-offs remain between specificity and clarity, and effective deployment will require robust content management, transparent validation pipelines, and integration within established clinical governance frameworks.

**Key points:**

- LLMs have comprehensive medical knowledge but require access to up-to-date, evidence-based, and locally relevant guidelines to be effective in CDS.
- Hallucinations (the generation of inaccurate or misleading information) remain a major limitation for LLMs in healthcare.
- Traditional information retrieval methods face several challenges in providing accurate, context-specific evidence.
- Retrieval-Augmented Generation (RAG) and graph-based RAG approaches have emerged as promising solutions to overcome these limitations.
- Renal medicine provides an ideal test domain to evaluate these models, given its complexity and reliance on nuanced, multidisciplinary decision-making.
- Studying LLM performance in kidney health can yield valuable insights into how such models can safely and effectively support complex clinical decision-making.

## Introduction

The modern healthcare landscape is becoming increasingly difficult to navigate - whilst waiting lists continue to grow ^1^, patients are more complex with multimorbidity and polypharmacy at an all-time high ^2^. Clinicians are expected to navigate electronic health records (EHRs), where patient data is incomplete or siloed and to stay up to date with an unprecedented volume of information, including vast repositories of biomedical literature, clinical research and guidelines ^3,4^. Clinicians often face challenges in synthesising vast amounts of information to make evidence-based decisions, a task complicated by time constraints and cognitive overload ^5,6^.

Clinical practice guidelines are essential for providing high-quality, standardised care. They condense extensive research into practical recommendations. However, their general nature often requires clinicians to invest considerable cognitive effort to interpret and apply them effectively in the specific context of each individual patient ^7^.

Traditional information retrieval systems can be effective for keyword-based searches but frequently fall short in providing nuanced, patient-specific insights derived from complex guidelines. They struggle to bridge the gap between generalised medical knowledge and the granular details embedded within a patient’s longitudinal health record, often leading to information fragmentation and potential delays in optimal care.

The emergence of LLMs holds transformative potential for healthcare, enabling sophisticated natural language understanding and generation ^8,9^. These models have demonstrated substantial medical knowledge, having passed medical exams ^10^, outperformed human clinicians in diagnosing complex cases ^11^, and recently even sequential questioning and diagnosis ^12^. The integration of LLMs into CDS systems is reshaping how clinicians access, interpret, and apply complex medical knowledge. However, significant limitations still prevent LLMs from being deployed in the real world, particularly in high-stakes environments such as healthcare; this includes technical limitations such as context window, hallucinations and biases ^13,14^, as well as knowledge limitations such as lack of real-world clinical data, knowledge cut-offs, and lack of hospital or region-specific guideline or protocol knowledge. Various approaches have been used to address knowledge limitations, including prompt engineering ^15^, medical domain fine-tuning ^16^ and the use of tools such as web search ^17^ with varying degrees of success. Among established techniques, RAG has demonstrated significant potential in enhancing LLMs by grounding outputs in curated knowledge sources, thereby improving clinical correctness and reducing hallucinations ^18,19,20^. In RAG architectures, external documents are retrieved in response to a user query and passed to a generation model, enabling more contextually relevant and trustworthy outputs, an essential capability in high-risk clinical environments.

Building on the foundations of RAG, GraphRAG is an advanced evolution designed for working with highly interconnected, structured information. Unlike standard RAG, which retrieves relevant passages from text, GraphRAG uses knowledge graphs that explicitly represent entities and their relationships, enabling more context-aware retrieval. It combines the precision of RAG in finding specific facts with the summarisation strengths of Query-Focused Summarisation (QFS), which captures broader themes of documents, tailored to the user’s question ^21^. This integration overcomes the limitations of each method: RAG alone struggles with complex, contextual queries, while QFS can be difficult to scale. GraphRAG therefore offers a scalable solution that can generate nuanced, comprehensive insights from large and diverse datasets ^22^. In healthcare, where patient data (e.g., diagnoses, medications, lab results) and medical knowledge (e.g., guideline recommendations, disease pathways) are inherently relational, knowledge graphs provide an ideal substrate for organising and querying information ^23^. GraphRAG has been used to enable more precise linking between patient features and relevant guideline logic, offering the potential for more personalised and explainable AI-assisted recommendations ^24,25^.

Managing CKD in primary care and other specialties outside of nephrology is challenging. Guidelines, which are based on biochemical markers and comorbidities, are difficult to align with individual patient data. While these guidelines condense extensive research into actionable recommendations, their generic nature demands significant cognitive effort from clinicians to accurately interpret and apply them to unique patient contexts. This can result in suboptimal adherence to evidence-based care and delayed interventions.

This paper examines the use of RAG and Graph RAG models for interpreting clinical guidelines, focusing on CKD as an example. We review the technical foundations of these models, assess their potential to improve adherence to guidelines and enhance decision-making, and discuss the factors to consider when implementing them in real-world clinical settings.

## Materials & Methodology

We evaluated 3 different CDS approaches (LLM, RAG, GraphRAG) on a dataset of 70 synthetic clinical CKD scenarios. Each scenario comprises a clinical vignette, demographic details, vital signs, and lab test results. Each CDS approach is evaluated on a set of nine questions per scenario that reflect real-world clinical practice.

**Figure 1.**
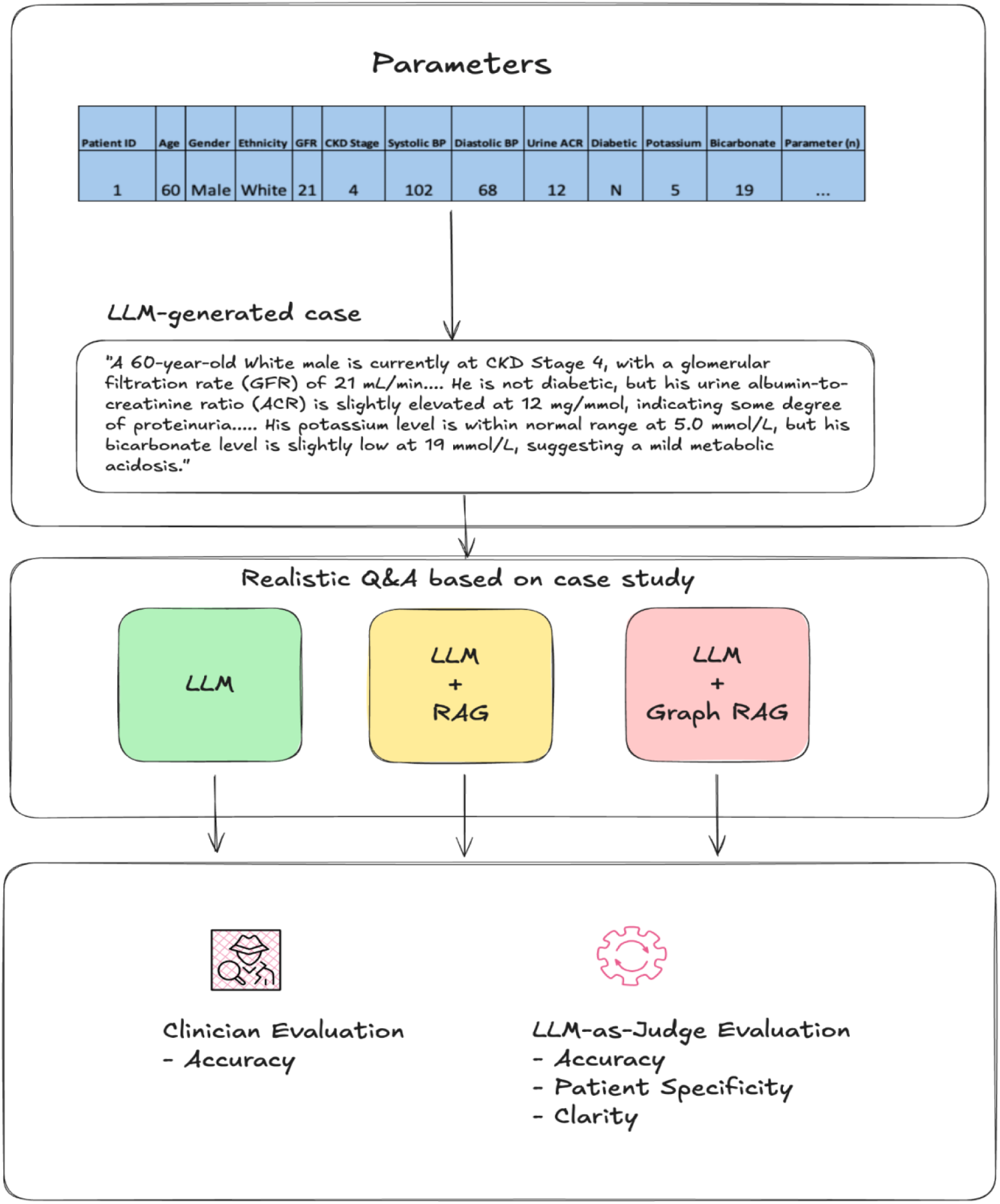
Summary of study design and evaluation methodology (LLM - Large Language model, RAG - Retrieval Augmented Generation)

### Data

We generated 70 synthetic CKD patients, each characterised by 17 features and a set of 9 clinician-curated question-answer pairs. These features include demographic information such as age, sex, and ethnicity, as well as clinical and laboratory data, including systolic and diastolic blood pressure, history of diabetes, glomerular filtration rate (GFR), potassium levels, bicarbonate levels, anemia status, bone profile, and urine albumin-to-creatinine ratio (ACR), ensuring that the patient scenarios are both diverse and realistic. A full list of features and their descriptions is available in the supplementary materials (Table 1). A qualified nephrologist created the 9 gold-standard question-answer pairs, and all recommendations are in keeping with the latest NICE CKD Guidelines^26^. These questions varied in format, including determinate, open-ended, and numerical (e.g., using the Kidney Failure Risk Equation, KFRE), to assess the models’ ability to generate accurate responses across different question types (Table 2).

Patient information was generated in tabular format; an example patient with corresponding features is presented in Table 1.

**Table 1:** Example patient (patient ID=1) from the list of synthetic patients.

| Patient ID | Age | Sex | Ethnicity | GFR | CKD Stage | Systolic BP | Diastolic BP | Urine ACR | Diabetic | Hgb (g/dL) | Ferritin (mcg/L) | Iron (µg/L) | Calcium (mmol/L) | Phosphate (mmol/L) | PTH (pg/mL) | Potassium (mmol/L) | Bicarbonate (mmol/L) |
| --- | --- | --- | --- | --- | --- | --- | --- | --- | --- | --- | --- | --- | --- | --- | --- | --- | --- |
| 1 | 60 | Male | White | 21 | 4 | 102 | 68 | 12 | N | 10.2 | 650 | 28 | 2.38 | 1.74 | 310 | 5 | 19 |

We used an LLM to convert each patient into a structured text describing all 17 features. An example patient description for the patient represented in Table 1 is as follows:

*“A 60-year-old White male is currently at CKD Stage 4, with a glomerular filtration rate (GFR) of 21 mL/min. His blood pressure is well controlled, with a systolic BP of 102 mmHg and a diastolic BP of 68 mmHg. He is not diabetic, but his urine albumin-to-creatinine ratio (ACR) is slightly elevated at 12 mg/mmol, indicating some degree of proteinuria. His haemoglobin level is 10.2 g/dL, suggesting mild anaemia, and his ferritin is elevated at 650 mcg/L. His iron level is relatively low at 28 µg/L. His calcium level is within normal range at 2.38 mmol/L, while his phosphate level is slightly elevated at 1.74 mmol/L. His parathyroid hormone (PTH) is significantly elevated at 310 pg/mL, indicating secondary hyperparathyroidism. His potassium level is within normal range at 5.0 mmol/L, but his bicarbonate level is slightly low at 19 mmol/L, suggesting a mild metabolic acidosis.”*

**Table 2.** List of questions for the synthetic CKD patients.

| Question Number | Archetype | Question |
| --- | --- | --- |
| 1 | Determinate task | Do I need to refer this patient to a nephrologist? |
| 2 | Determinate task | What is the minimum frequency I need to check kidney function in this patient? |
| 3 | Determinate task | What should I do about this patient's blood pressure? If it needs treatment, what is the best first option? |
| 4 | Open-ended task | How do I manage anaemia in this patient? |
| 5 | Open-ended task | How do I manage bone disease in this patient? |
| 6 | Open-ended task | What is the appropriate clinical action regarding this patient's potassium levels? |
| 7 | Open-ended task | What is the appropriate clinical action regarding this patient's bicarbonate levels? |
| 8 | Enumerative/creative | What other medication treatment options should I consider for their CKD? |
| 9 | Numerical formula | What is the risk of end-stage kidney disease in this patient in 5 years using the Kidney Failure Risk Index (KFRE)? |

### Methodology

We evaluated three popular LLM architectures:

(1) Out-of-the-box LLM without specific knowledge of CKD clinical guidelines (LLM)
(2) LLM with RAG of NICE CKD clinical guidelines (LLM + RAG)
(3) LLM with RAG further enhanced by a knowledge graph generated via the Graph-RAG algorithm. (LLM + graph RAG)

All experiments were run in Python 3.10. We used ChatGPT-4o, provided by Azure OpenAI, as our LLM choice in this study. We implemented RAG and GraphRAG for models 2 & 3 based on the Python llama_index package. The prompts and further model information are described in the supplementary materials.

### Baseline LLM Model (No CKD-Specific Knowledge)

In this experiment, the LLM was provided only with the synthetic patient data and prompted to generate recommendations based on its general knowledge. We presented the LLM with a prompt for each patient that included two input/output examples, the patient description (as illustrated in the Data section), and all nine questions in a single prompt.

### LLM with RAG on Vector-Indexed NICE CKD Guidelines

Here, we linked the LLM to the NICE CKD clinical guidelines using a vector-indexed RAG pipeline built with the Python *llama_index* package. The guidelines were split into small sections and converted into vector embeddings so that, for each patient question, the system could retrieve the most relevant guideline content based on similarity. For each question, the RAG system returned the top five most relevant guideline sections (“chunks”), which were then added to the prompt to provide the LLM with a focused clinical context. Because each question required different guideline information, we generated nine separate prompts per patient, each enriched with its own targeted RAG-retrieved context.

### LLM with Graph-RAG on NICE CKD Guidelines

We employed a more advanced RAG technique, incorporating a knowledge graph (GraphRAG) based on the NICE CKD guidelines. This knowledge graph was constructed using the original MS GraphRag implementation ^27^, designed to map complex relationships between different aspects of the CKD guidelines, such as symptoms, stages, and recommended treatments. By using this knowledge graph, the LLM could access and interpret multi-layered, interconnected information from the guidelines, leading to a more sophisticated retrieval mechanism. This approach aimed to evaluate if the structured, relational information within the knowledge graph could further enhance the accuracy of the model’s management recommendations for synthetic patient scenarios. Similar to vector-indexed RAG experiments, we separately queried each question for a given patient in GraphRAG experiments, as combining all contexts and questions into a single prompt diluted relevance and often exceeded token limits.

Details of the prompts used for each model are described in the supplementary materials.

### Evaluation Metrics

We assessed each CDS pipeline using four evaluation metrics: one clinician assessment, considered the gold standard, and four using LLM-as-judge metrics (GPT-4o, Azure OpenAI, June 2025 model snapshot).

#### Clinical Correctness – Clinician Assessment

A consultant nephrologist, EA, independently judged every model response. For each vignette, the clinician reviewed the patient context and the candidate answer and assigned a binary label: **Correct** (1) when the response was clinically sound, actionable, and free of material errors; **Incorrect** (0) when it contained critical omissions or factual inaccuracies. This manual label serves as the primary ground-truth outcome against which all automated metrics are compared.

#### Clinical Correctness - LLM

We used an LLM-as-judge to compare every submitted answer with the curated reference answer. The prompt contained the original question, the patient identifier, both answers, and an instruction to decide whether the two conveyed the same essential clinical content. The LLM responded with **TRUE** when the answers were semantically equivalent, and with **FALSE** when it detected material omissions, contradictions, or errors, accompanied by a concise explanatory note.

#### Patient-Specificity

We next measured the degree to which responses were tailored to the individual case. The evaluation LLM received the patient demographics and laboratory profile, along with the answer. It was instructed to count how many of the 17 predefined patient attributes, including age, sex, ethnicity, kidney function, blood pressure readings, metabolic parameters, and anaemia markers, were explicitly mentioned or clearly incorporated into the response. The model returned both the total count and the list of identified attributes for each question. For reporting purposes, the number of relevant features returned was standardised by the specific question number. This allowed for a fair comparison of how well each methodology tailored its response to the patient characteristics based on the specific question.

#### Clarity

We used LLM-as-judge to rate each answer’s clarity and conciseness on a scale from 1 (excessively verbose or padded) to 10 (perfectly concise, zero redundancy). The prompt framed the task from a clinical reader’s perspective, explicitly instructing the judge not to penalise accurate medical terminology or standard clinical abbreviations, but rather to focus on unnecessary verbosity, repetition, or poor organisation.

## Results

### NICE CKD Guidelines Knowledge Graph

Using the out-of-the-box MS GraphRAG pipeline, we ingested the full text of the NICE CKD guidelines and obtained the knowledge graph that we subsequently explored in Gephi ^28^. A structured map showing how different concepts within the document are connected. The resulting graph contained 208 unique terms (nodes) linked by 280 relationships (edges). On average, each concept was connected to just over one other concept, and less than 1% of all possible links were present. In other words, the guideline’s structure is quite sparse. This reflects NICE’s modular approach to presenting its recommendations, as discrete statements about investigations, treatments, and follow-up, rather than as a tightly interconnected web of biomedical terminology, as in large clinical ontologies.

Figure 2 shows a two-dimensional visualisation of this CKD knowledge graph. ^29^ The layout was generated using a standard algorithm that positions closely related terms closer together. We then identified clusters of related terms (communities) using a modularity detection method in Gephi, which coloured each cluster differently.^30^ Three major clusters were identified:

- a pink hub centred on the term *NICE*
- a green hub around *CKD*
- a blue hub around *Chronic Kidney Disease*.

**Figure 2.**
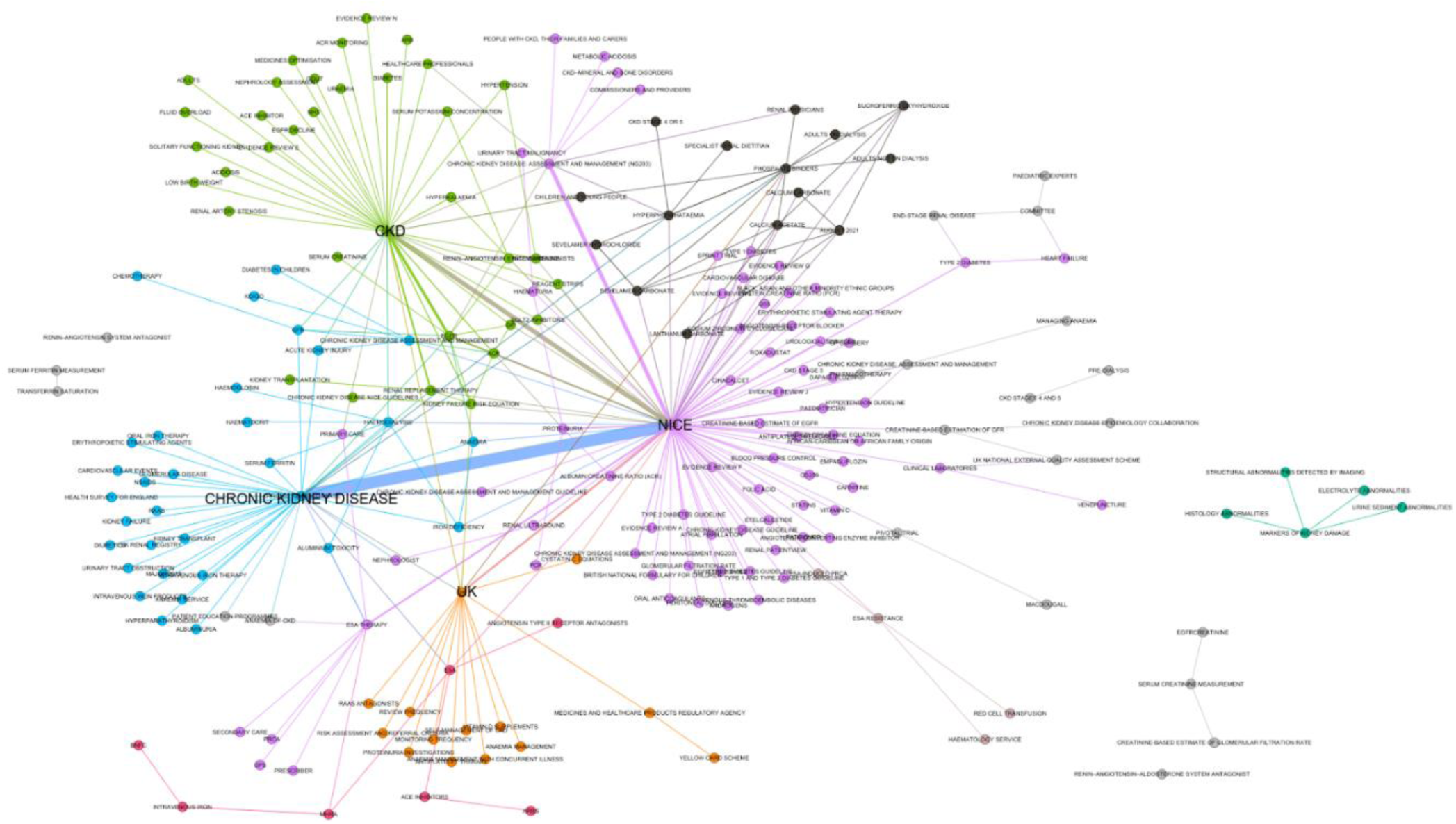
Two-dimensional layout of the CKD knowledge Graph

Together, these three hubs accounted for nearly half of all terms in the graph. From a structural point of view, *NICE* acts as the main connector or “super-hub,” linking almost every theme that runs through the guideline. The terms *CKD* and *Chronic Kidney Disease* form the next level of connectivity, while key clinical concepts such as *eGFR*, *albumin-to-creatinine ratio*, and *phosphate binders* form smaller but distinct clusters. The links between these concepts mostly represent clinical actions, for example, “used,” “provides,” “treatment,” or “monitoring,” typically link a NICE recommendation to a test or therapy.

### Comparing The Three CDS Approaches

We evaluated three pipelines—LLM, LLM + RAG, and LLM + GraphRAG—on 70 synthetic CKD cases with 17 structured clinical features and 9 decision-support questions. For every patient-question pair, we calculated clinical correctness, clarity, and patient-specificity as defined in the evaluation metrics section.

To aid interpretation, we grouped the nine evaluation questions into six archetypes that mirror the type of clinical reasoning required to answer the question accurately. These are shown in Table 3.

**Table 3.** Question archetypes and cognitive demand on the model for answering.

| Archetype | Example vignette (Q-no) | Cognitive demand |
| --- | --- | --- |
| Binary rule-out / referral | Nephrology referral? (Q 1) | Apply a single threshold |
| Structured look-up/scheduling | Minimum monitoring frequency (Q 2) | Retrieve table entry |
| Algorithmic choice | First-line antihypertensive (Q 3) | Follow the decision tree |
| Open-ended management plan | Anaemia, bone, potassium, bicarbonate (Q 4-7) | Synthesise a multi-step plan |
| Extra-guideline drug expansion | Additional CKD drugs (Q 8) | Draw on external knowledge beyond the indexed guideline |
| Numerical formula | 5-yr KFRE risk (Q 9) | Perform multi-step maths |

#### Clinical Correctness - Clinician assessment

The overall clinical correctness, as judged by the clinician, favours the retrieval pipelines as follows: LLM +RAG scores 76.6%, LLM + GraphRAG scores 75.6%, and LLM scores 70.8%. When breaking down correctness by question, GraphRAG performs best on questions 1, 5, and 9; Vector-RAG excels on questions 2 and 7; while LLM performs best on questions 3, 4, 6, and 8. Table 4 shows the clinical correctness scores for each question and model.

**Table 4.** Comparison of the correctness of each question with answers judged by a clinician (including archetype tags).

| Q-no | Archetype | Clinical task | LLM | LLM + RAG | LLM + GraphRAG |
| --- | --- | --- | --- | --- | --- |
| 1 | Determinate task | Referral decision | 0.74 | 0.81 | **0.82** |
| 2 | Determinate task | Monitoring interval | 0.44 | **1** | **1** |
| 3 | Determinate task | First-line antihypertensive | **0.93** | 0.89 | 0.89 |
| 4 | Open-ended task | Anaemia management | **0.86** | 0.8 | 0.84 |
| 5 | Open-ended task | Bone disease plan | 0.45 | 0.48 | **0.48** |
| 6 | Open-ended task | Potassium threshold | **0.97** | 0.93 | 0.9 |
| 7 | Open-ended task | Bicarbonate threshold | 0.86 | **1** | 0.97 |
| 8 | Extra-guideline drug expansion | Extra-guideline drug expansion | **1** | 0.28 | 0.1 |
| 9 | Numerical formula | 5-yr KFRE risk | 0.03 | 0.28 | **0.31** |

For the binary referral task (Q1), LLM + GraphRAG slightly outperformed the others, scoring 82% compared to 81% for LLM + RAG and 74% for the LLM model. This suggests a benefit from incorporating contextual features beyond a single threshold. Across the open-ended management plans (Q4–7), retrieval methods had a slight overall advantage (≈ 0.80) over the LLM model (≈ 0.78), although the bone disease task (Q5) proved uniformly challenging across all methods (scores ranged from ≈ 0.45 to 0.48). In the extra-guideline drug expansion task (Q8), the LLM scored 100%, whereas the LLM + RAG variants performed poorly because the indexed NICE guideline lacked key drug classes, notably SGLT-2 inhibitors and finerenone. For the numerical formula task (Q9, KFRE), LLM + GraphRAG achieved the highest performance at 31%, followed by LLM + RAG at 28%, while the LLM model scored only 3%.

To complement the question-level results in Table 4, we aggregated clinician-judged correctness by archetype and report the corresponding means and standard deviations in Table 5. As expected, determinate tasks showed low variability across methods, whereas open-ended questions showed greater variability, reflecting broader clinical reasoning demands.

**Table 5.** Clinician-judged clinical correctness by methodology with associated archetype.

| Archetype | LLM | LLM + RAG | LLM + GraphRAG | # Questions |
| --- | --- | --- | --- | --- |
| Determinate task | $0.7 \pm 0.2$ | **$0.9 \pm 0.1$** | **$0.9 \pm 0.1$** | 3 |
| Open-ended task | **0.79** ± 0.2 | 0.8 ± 0.22 | 0.80 ± 0.2 | 4 |
| Extra-guideline drug expansion | **1** ± 0 | 0.28 ± 0 | 0.1 ± 0 | 1 |
| Numerical formula | 0.03 ± 0 | 0.28 ± 0 | **0.31** ± 0 | 1 |

#### Clinical Correctness — LLM-as-judge

With LLM-based judging, LLM + GraphRAG either leads or shares the top position in six out of the nine clinical questions (Q2-4, 7, and 9) and closely follows LLM + RAG in Q1. However, its overall micro-correctness metric seems average because performance on Q8 (regarding "additional CKD drugs") declines significantly for both RAG pipelines. The details of this pattern are in Table 6.

**Table 6.** Evaluation of clinical correctness per question, assessed by LLM, with associated archetype tag.

| Q-no | Archetype | Clinical task | LLM | LLM + RAG | LLM + GraphRAG |
| --- | --- | --- | --- | --- | --- |
| 1 | Determinate task | Referral decision | 0.70 | **0.82** | 0.78 |
| 2 | Determinate task | Monitoring interval | 0.93 | **1.00** | **1.00** |
| 3 | Determinate task | First-line antihypertensive | 0.79 | 0.89 | **0.93** |
| 4 | Open-ended task | Anaemia management | 0.38 | 0.16 | **0.65** |
| 5 | Open-ended task | Bone disease plan | **0.24** | 0.16 | 0.18 |
| 6 | Open-ended task | Potassium threshold | **0.93** | 0.71 | 0.33 |
| 7 | Open-ended task | Bicarbonate threshold | 0.82 | 0.78 | **0.93** |
| 8 | Extra-guideline drug expansion | Extra-guideline drug expansion | **0.62** | 0.04 | 0.02 |
| 9 | Numerical formula | 5-yr KFRE risk | 0.02 | 0.02 | **0.05** |

For KFRE (Q 9), all methods struggle because the formula is embedded as an image in the guideline. Despite our effort of processing the image via the LLM (ChatGPT-4o) and including the result in the corpus without explicit function-calling the pipelines incorrectly apply or guess coefficients, leading to very low absolute clinical correctness (GraphRAG 0.05 vs 0.02 for others). At the archetype level, accuracies decline with cognitive complexity: structured look-ups yield the highest accuracies, followed by algorithmic choices. Open-ended tasks and extra-guideline tasks have lower accuracies, while numerical formulae yield the lowest results. The results are shown in Table 7.

**Table 7.** LLM-judged clinical correctness by methodology with associated archetype.

| Archetype | LLM | LLM + RAG | LLM + GraphRAG | # Questions |
| --- | --- | --- | --- | --- |
| Determinate task | 0.81 ± 0.02 | **0.9 ± 0.03** | **0.9 ± 0.03** | 3 |
| Open-ended task | **0.59** ± 0.1 | 0.45 ± 0.11 | 0.52 ± 0.13 | 4 |
| Extra-guideline drug expansion | **0.62** ± 0 | 0.04 ± 0 | 0.02 ± 0 | 1 |
| Numerical formula | 0.02 ± 0 | 0.02 ± 0 | **0.05** ± 0 | 1 |

Across all nine questions, LLM achieves ≈ 60%, LLM + GraphRAG ≈ 54%, LLM + RAG ≈ 51%. Excluding Q 8, LLM+ GraphRAG edges ahead at ≈ 61% (LLM ≈ 60%, LLM + RAG ≈ 57%). The percentages indicate the proportion of patient–question pairs judged correct within each category.

The LLM-based results mirror the clinician-based trends while highlighting two additional points. First, retrieval is most effective when the answer is directly present in the guideline (referral criteria, monitoring cadence, ACE-i/ARB choice; Q 1–3). GraphRAG adds useful context (e.g., integrating urine-ACR into antihypertensive advice and propagating laboratory values into anaemia plans), which explains its advantage in Q 3–4. Secondly, the LLM model performs best when questions require expansion beyond the indexed guideline (Q 8) or a pragmatic single threshold (Q 6). The Q8 deficit for RAG variants is a scope artefact rather than a reasoning flaw: key classes such as SGLT-2 inhibitors and finerenone are absent from the indexed NICE CKD guideline, so faithful retrieval omits them.

### Shared observations (clinician & LLM)

We observe that across both clinician and LLM judging, three themes recur. Retrieval excels for structured look-ups such as monitoring intervals (Q 2), where both RAG and graph-based pipelines reach 100% under clinician scoring and 100% under automated judging. The LLM model is strongest when answers must reach beyond the indexed guideline or rely on a single pragmatic threshold; this explains its leadership on extra-guideline drug expansion (Q 8) and potassium thresholds (Q 6). GraphRAG is relatively strongest on KFRE (Q 9), although absolute accuracies remain low across methods because the risk equation is only present as an image in the guideline.

The deficit for RAG on Q 8 is best interpreted as a document-scope artefact, not a failure of enumerative reasoning: important drug classes (notably SGLT-2 inhibitors and finerenone) are not contained in the single NICE CKD document indexed by the retrieval pipelines, so faithful retrieval necessarily omits them. Finally, overall rankings diverge by judge: clinician-judged results rate both retrieval pipelines higher overall than the LLM model (≈ 76–76% vs 71%), whereas the LLM-as-judge ranks the LLM model highest (≈ 60% vs 54% and 51%). In absolute terms, the LLM-based judge under-calls clinical acceptability by ∼12–27 percentage points (LLM −12 pp; LLM + RAG −27 pp; LLM + GraphRAG −22 pp).

#### Clarity and patient-specificity

Table 8 shows the results for clarity and patient-specificity across methods. GraphRAG scores lowest in clarity because its graph walks often return long guideline excerpts, which add unnecessary length and obscure the key recommendation. A brief post-processing step to trim these could improve clarity by about 1.5 points. On the other hand, GraphRAG performs best in terms of patient-specificity, as it consistently integrates individual lab values (such as urine ACR and ferritin) into its responses, especially noticeable in Questions 3, 4, and 7.

**Table 8.** Clarity and patient specificity of responses using different models.

| Metric | LLM | LLM + RAG | LLM + GraphRAG |
| --- | --- | --- | --- |
| Clarity (↑ better) | **$9.01 \pm 0.45$** | $8.53 \pm 0.52$ | $7.80 \pm 0.47$ |
| Patient-specificity | $2.35 \pm 0.88$ | $3.77 \pm 1.52$ | **$6.59 \pm 2.47$** |

Table 9 shows that this trade-off is highly dependent on question type: GraphRAG offers the richest patient contextualisation in open-ended management questions, but at the cost of marked clarity, while the three models behave far more similarly in determinate guideline look-ups (Q1–3). Notably, guideline-expansion questions (Q8) show high specificity but limited clarity, reflecting the broader clinical knowledge required and the constraints of the retrieval corpus.

**Table 9.** Clarity and patient specificity of responses using different models, aggregated by question archetype.

| Category | Metric | LLM | LLM + RAG | LLM + GraphRAG |
| --- | --- | --- | --- | --- |
| **Determinate (Q1-3)** | **Clarity** | 9.27 ± 0.46 | 8.92 ± 0.29 | 7.95 ± 0.35 |
|  | **Specificity** | 2.62 ± 0.72 | 3.65 ± 1.29 | 6.04 ± 1.81 |
| **Open-ended (Q4-7)** | **Clarity** | 9.00 ± 0.18 | 8.48 ± 0.53 | 7.63 ± 0.54 |
|  | **Specificity** | 1.99 ± 0.90 | 3.13 ± 1.06 | 7.47 ± 2.86 |
| **Guideline expansion (Q8)** | **Clarity** | 8.93 ± 0.26 | 8.11 ± 0.31 | 8.00 ± 0.00 |
|  | **Specificity** | 2.36 ± 0.49 | 6.44 ± 1.58 | 7.33 ± 1.20 |
| **Numerical formula (Q9)** | **Clarity** | 8.35 ± 0.55 | 7.96 ± 0.19 | 7.84 ± 0.50 |
|  | **Specificity** | 2.98 ± 0.89 | 4.00 ± 0.00 | 4.00 ± 0.00 |

## Discussion

This study demonstrates the feasibility and limitations of applying RAG and GraphRAG to deliver guideline-aligned, patient-specific recommendations in the management of CKD. Using a synthetic cohort of 70 patients and 9 clinically relevant questions derived from NICE guidance, we evaluated three models: a baseline LLM, a RAG pipeline using vector search, and a GraphRAG pipeline. Our findings yield several key insights into performance patterns, use-case boundaries, and opportunities for future refinement.

### Matching Pipeline to Clinical Task Type

Performance varied markedly across question types. Where the guideline offered discrete thresholds or tabular entries (e.g., referral criteria, monitoring frequency), RAG-based models achieved perfect or near-perfect clinical correctness. GraphRAG further improved performance in algorithmic decisions, such as choosing a first-line antihypertensive, by retrieving semantically related context nodes (e.g., integrating urine ACR and blood pressure into treatment logic).

Clinician scoring confirmed these trends: RAG pipelines consistently outperformed the baseline on monitoring and referral questions (Q1–2), but showed less advantage on threshold-based tasks (Q3, Q6–7). By contrast, the LLM-judge rated GraphRAG more highly on open-ended synthesis tasks (Q4, Q7), yet penalised it on potassium management (Q6). These differences suggest that tasks involving clear thresholds or table look-ups are relatively stable across evaluators, whereas complex management plans are more sensitive to whether the assessor is human or model-based.

These findings are consistent with recent research demonstrating that custom LLM models grounded in evidence can significantly improve clinical correctness for evidence-based medicine ^31^. Conversely, for open-ended synthesis tasks such as anaemia or bicarbonate management, GraphRAG outperformed both the baseline and RAG, indicating its potential to support complex reasoning by chaining indirect relationships across the knowledge graph.

In contrast, the baseline LLM (GPT-4o) excelled at tasks requiring either knowledge beyond the indexed NICE document (e.g., listing additional CKD therapies in Q8) or simple one-line numerical thresholds embedded in general medical training (e.g., potassium cut-offs in Q6). This shows that baseline models may outperform retrieval-based systems when (a) key content is absent from the indexed corpus, or (b) guideline fidelity is less important than broad clinical recall.

### Role of Guideline Scope and Content Gaps

The retrieval-based systems performed poorly when the guideline lacked explicit coverage of the clinical issue, as in Q8. Here, both RAG models failed to recommend SGLT-2 inhibitors or finerenone, as these drug classes were not present in the indexed NICE CKD guideline. This limitation underscores a key deployment consideration: while RAG systems can improve guideline fidelity, they are inherently constrained by the scope of the input corpus. In real-world settings, pipeline performance will depend heavily on the breadth, recency, and granularity of the indexed guidance. Practically, this suggests expanding a target corpus to include drug-class guidance and other dependencies for the tasks that require “beyond-guideline” information.

### Computation and Format Challenges

KFRE estimation (Q9) highlighted a persistent challenge for both LLM and RAG methods: structured information, such as equations or tables embedded as images, is poorly ingested. Even after converting the KFRE equation image into text, LLMs produced inconsistent outputs and hallucinated coefficients. GraphRAG occasionally approximated the correct formula through deeper graph exploration, but clinical correctness remained low across all methods. This suggests that hybrid approaches combining LLMs with symbolic calculators or structured tables may be required to handle guideline elements expressed in non-textual form. Adding OCR (Optical Character Recognition) or a function-calling step would likely improve performance.

### Trade-offs Between Clarity and Specificity

GraphRAG produced the most patient-specific outputs, consistently integrating laboratory values and demographic details into its recommendations. This strength, however, came at the expense of clarity, as responses were often lengthy and less concise due to the verbose guideline excerpts retrieved during graph traversal. Introducing post-generation filtering or summarisation could improve readability and make outputs more accessible for time-pressured clinicians.

### Practical Implications and Validation

The modular design of RAG pipelines makes them well-suited for local customisation. Hospitals can swap NICE guidance for local trust protocols, and adaptation to new specialties or jurisdictions is straightforward. This contrasts with fine-tuned LLMs, which require retraining for each guideline variant. However, this flexibility must be matched with rigorous validation pipelines that are monitored and updated regularly, particularly when used for clinical decision support. Platforms such as [Valmed.ai](https://profvalmed.com/) and related efforts exploring retrieval and validation pipelines demonstrate a growing recognition that robust retrieval and auditable validation are essential to bring AI to the bedside ^23,32^.

### Improving GraphRAG with Ontology Refinement

While the knowledge graph already adds value by linking context across multiple steps, further improvements could be achieved through lightweight ontology processing. At present, lexical duplicates (e.g., *‘CKD’* vs *‘Chronic Kidney Disease’*) fragment the graph and weaken query coverage, while inconsistent edge labels (e.g., *‘guideline recommends’* vs *‘guideline provides’*) reduce path consistency. Adding a normalisation layer based on UMLS (Unified Medical Language System) or SNOMED, combined with predicate standardisation and a simple class–instance hierarchy, would minimise redundancy, improve node connectivity, and surface more coherent evidence chains, without altering the underlying pipeline.

### Divergence Between Clinician and LLM Judgements

A key finding was that performance rankings differed between clinician-judged accuracy and LLM-judged accuracy. For determinate tasks, both judges generally favoured RAG pipelines, but the extent of their preferences varied. For the monitoring interval (Q2), the clinician gave the baseline model a low score of 0.44, while the LLM judge rated it highly at 0.93.

In open-ended tasks, the clinician tended to give higher scores overall. In contrast, the LLM judge sharply penalised the baseline and RAG models in the context of anaemia and bone disease. Regarding potassium thresholds (Q6), the clinician rated all models nearly perfectly, whereas the LLM judge scored GraphRAG significantly lower at 0.33.

Finally, for creative and formula-based questions (Q8–9), the clinician recognised partial retrieval benefits that the LLM judge largely overlooked. This divergence highlights that evaluation outcomes depend strongly on the perspective of the judge.

Several limitations of this work warrant consideration. First, all experiments were performed on synthetic patient profiles, which, while constructed to be diverse and clinically plausible, cannot capture the full variability and complexity of real-world practice. Second, we deliberately used the out-of-the-box GraphRAG package as released by its authors, without additional ontology refinement, disambiguation, or predicate canonicalisation. This choice was intentional to evaluate performance in its default form, as it would be encountered by clinical researchers or practitioners adopting the methodology directly. The limitations with this approach included:

- Duplicate terms: the system treats different spellings or abbreviations (for example, *CKD* and *chronic kidney disease*) as separate entities, which unnecessarily fragments the graph and reduces its usefulness
- Inconsistent relationship labels: because the relationships are extracted directly from the text, similar phrases such as “guideline recommends,” “guideline offers,” or “guideline provides” are treated as distinct, even though they mean the same thing. This makes it harder to search for related recommendations.
- Lack of hierarchical structure: the current system represents all terms at the same level. As a result, broad categories (such as *Anaemia*) appear alongside specific examples (like *calcium acetate*), which prevents more sophisticated reasoning about classes of conditions or treatments.

Third, we restricted our corpus to a single guideline (NICE CKD) and did not expand the context to other related domains, such as the hypertension guideline. The content and structure of the underlying guideline corpus inherently constrain the performance of RAG-based systems. When key treatments or calculations are missing (e.g., SGLT-2 inhibitors, KFRE formula), even a flawless retrieval strategy yields incomplete or outdated recommendations. These constraints mean that our findings should be interpreted as an early feasibility signal rather than definitive evidence of clinical utility.

## Conclusion

This study demonstrates the potential of RAG and GraphRAG to support safe, guideline-aligned decision-making in CKD care. By grounding LLM outputs in curated clinical guidance, RAG methods significantly improved clinical correctness, patient specificity, and adherence to structured protocols, especially in decision types governed by thresholds, tables, or algorithmic rules.

GraphRAG extended these capabilities further by capturing relational context, enabling reasoning across interconnected guideline elements. This proved particularly useful in complex, open-ended management tasks such as anaemia or bicarbonate treatment, where traditional retrieval alone was insufficient.

In summary, RAG and GraphRAG provide a scalable, auditable foundation for LLM-based clinical decision support, especially in settings that demand fidelity to evolving standards of care. To fully realise their promise, these systems must be paired with robust content management, evaluation workflows, and ongoing alignment with clinical governance frameworks.

## Data Availability

All data produced in the present study are available upon reasonable request to the authors

## Supplementary Materials

Table 1 lists the patient features and their descriptions used in the study.

**Table 1:**
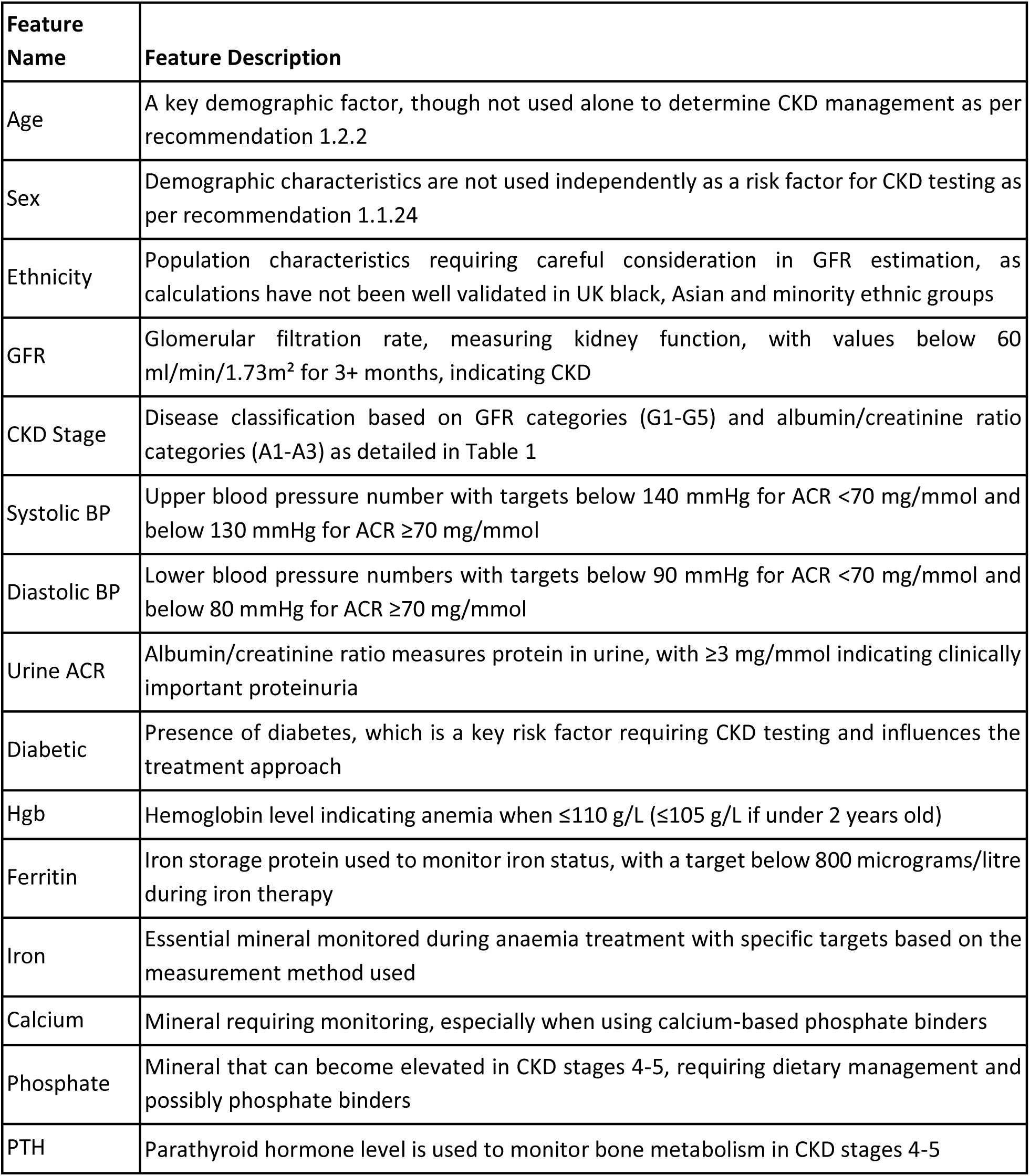
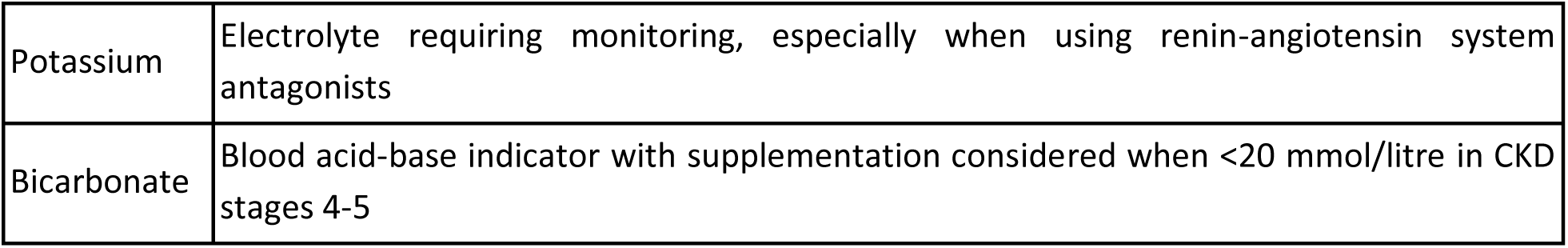
Patient features and their description.

| Feature Name | Feature Description |
| --- | --- |
| Age | A key demographic factor, though not used alone to determine CKD management as per recommendation 1.2.2 |
| Sex | Demographic characteristics are not used independently as a risk factor for CKD testing as per recommendation 1.1.24 |
| Ethnicity | Population characteristics requiring careful consideration in GFR estimation, as calculations have not been well validated in UK black, Asian and minority ethnic groups |
| GFR | Glomerular filtration rate, measuring kidney function, with values below 60 ml/min/1.73m<sup>2</sup> for 3+ months, indicating CKD |
| CKD Stage | Disease classification based on GFR categories (G1-G5) and albumin/creatinine ratio categories (A1-A3) as detailed in Table 1 |
| Systolic BP | Upper blood pressure number with targets below 140 mmHg for ACR <70 mg/mmol and below 130 mmHg for ACR ≥70 mg/mmol |
| Diastolic BP | Lower blood pressure numbers with targets below 90 mmHg for ACR <70 mg/mmol and below 80 mmHg for ACR ≥70 mg/mmol |
| Urine ACR | Albumin/creatinine ratio measures protein in urine, with ≥3 mg/mmol indicating clinically important proteinuria |
| Diabetic | Presence of diabetes, which is a key risk factor requiring CKD testing and influences the treatment approach |
| Hgb | Hemoglobin level indicating anemia when ≤110 g/L (≤105 g/L if under 2 years old) |
| Ferritin | Iron storage protein used to monitor iron status, with a target below 800 micrograms/litre during iron therapy |
| Iron | Essential mineral monitored during anaemia treatment with specific targets based on the measurement method used |
| Calcium | Mineral requiring monitoring, especially when using calcium-based phosphate binders |
| Phosphate | Mineral that can become elevated in CKD stages 4-5, requiring dietary management and possibly phosphate binders |
| PTH | Parathyroid hormone level is used to monitor bone metabolism in CKD stages 4-5 |
| Potassium | Electrolyte requiring monitoring, especially when using renin-angiotensin system antagonists |
| Bicarbonate | Blood acid-base indicator with supplementation considered when <20 mmol/litre in CKD stages 4-5 |

### Considerations about open-ended questions

Some questions had to be iterated as they were flagged by the content filters of the LLMs. For instance, Question 7 was originally phrased as “What should I do about bicarbonate?”, which triggered the LLM’s **"self-harm" content filter trigger** (’self_harm’: {’filtered’: True, ‘severity’: ‘high’}), possibly due to ambiguity and the model (or filter) misreading it as a general question about ingesting bicarbonate (which could have toxicity concerns). The question was rephrased to its current form in Table 2.

### Prompt Structure for different models

Prompt structure for LLM (GPT4o):

~~~
patient_specific_query = f"""You are an AI assistant that helps in answering clinical questions..
For this specific patient with key patient parameters:
{patient_params_text}
Please answer these questions: Q1: {question_1_text}, Q2: {question_2_text},…, Q9:
{question_9_text}
"""
~~~

Prompt Structure for RAG:

~~~
question_prompt = f"""You are an AI assistant that helps analyse patient cases based on clinical guidelines.
Answer this question concisely: {q_text} for this specific patient with key patient parameters:
{patient_params_text}
Based on the following NICE guideline information:
{context}
"""
~~~

Prompt Structure for GraphRAG:

~~~
patient_specific_query = f"""You are an AI assistant that helps analyse patient cases based on clinical guidelines.
Answer this question concisely: {q_text} for this specific patient with key patient parameters:
{patient_params_text}
"""
~~~

We used the local search for GraphRAG to answer the patient questions, as these are targeted and focused on specific aspects of CKD management (referrals, monitoring frequency, blood pressure management, anaemia, bone disease, electrolyte management, etc.). Specifically, a community level parameter of 2 for local search in order to provide the right balance of specificity, focusing on CKD management guidelines while maintaining enough context about related conditions and treatments (note the local search contrasts with global search, which is more appropriate for answering questions like “What are all the major considerations in CKD management?”. This approach is more resource-intensive because it analyses the entire knowledge graph to identify patterns, relationships, and overarching themes.

## References

1. Zaranko, M. W. B. The past and future of NHS waiting lists in England. https://ifs.org.uk/publications/past-and-future-nhs-waiting-lists-england (Published on 29 February 2024).

2. Chowdhury, S. R., Chandra Das, D., Sunna, T. C., Beyene, J. & Hossain, A. Global and regional prevalence of multimorbidity in the adult population in community settings: a systematic review and meta-analysis. EClinicalMedicine 57, 101860 (2023).

3. Holmes, J. H. et al. Why is the electronic health record so challenging for research and clinical care? Methods Inf. Med. 60, 32–48 (2021).

4. Kim, M. K., Rouphael, C., McMichael, J., Welch, N. & Dasarathy, S. Challenges in and opportunities for electronic health record-based data analysis and interpretation. Gut Liver 18, 201–208 (2024).

5. Klerings, I., Weinhandl, A. S. & Thaler, K. J. Information overload in healthcare: too much of a good thing? Z. Evid. Fortbild. Qual. Gesundhwes. 109, 285–290 (2015).

6. Sbaffi, L., Walton, J., Blenkinsopp, J. & Walton, G. Information overload in emergency medicine physicians: A multisite case study exploring the causes, impact, and solutions in four north England National Health Service trusts. J. Med. Internet Res. 22, e19126 (2020).

7. Guerra-Farfan, E. et al. Clinical practice guidelines: The good, the bad, and the ugly. Injury 54 **Suppl 3**, S26–S29 (2023).

8. Nazi, Z. A. & Peng, W. Large language models in healthcare and medical domain: A review. arXiv [cs.CL] (2024) doi:10.48550/ARXIV.2401.06775.

9. Yu, E. et al. Large language models in medicine: Applications, challenges, and future directions. Int. J. Med. Sci. 22, 2792–2801 (2025).

10. Kung, T. H. et al. Performance of ChatGPT on USMLE: Potential for AI-assisted medical education using large language models. PLOS Digital Health 2, e0000198 (2023).

11. McDuff, D. et al. Towards accurate differential diagnosis with large language models. Nature 642, 451–457 (2025).

12. Nori, H., et al. Sequential Diagnosis with Language Models. (2025).

13. Xu, Z., Jain, S. & Kankanhalli, M. Hallucination is inevitable: An innate limitation of large language models. arXiv [cs.CL] (2024).

14. Au Yeung, J., et al. AI chatbots not yet ready for clinical use. Front. Digit. Health 5, 1161098 (2023).

15. Wang, L. et al. Prompt engineering in consistency and reliability with the evidence-based guideline for LLMs. npj Digital Medicine 7, 1–9 (2024).

16. Singhal, K. et al. Toward expert-level medical question answering with large language models. Nature Medicine 31, 943–950 (2025).

17. Shen, Y., et al. HuggingGPT: Solving AI Tasks with ChatGPT and its Friends in Hugging Face. (2023).

18. Yang, R. et al. Retrieval-augmented generation for generative artificial intelligence in health care. Npj Health Syst. 2, (2025).

19. Lewis, P., et al. Retrieval-augmented generation for knowledge-intensive NLP tasks. arXiv [cs.CL] (2020) doi:10.48550/ARXIV.2005.11401.

20. Semnani, S. J., Yao, V. Z., Zhang, H. C. & Lam, M. S. WikiChat: Stopping the hallucination of large language model chatbots by few-shot grounding on Wikipedia. arXiv [cs.CL] (2023).

21. Edge, D. et al. From local to global: A graph RAG approach to query-focused summarization. arXiv [cs.CL] (2024).

22. Hu, Y., et al. GRAG: Graph Retrieval-Augmented Generation. arXiv [cs.LG] (2025).

23. Wu, J. et al. Medical graph RAG: Towards safe medical Large Language Model via graph retrieval-Augmented Generation. arXiv [cs.CV] (2024) doi:10.48550/ARXIV.2408.04187.

24. Madrid-García, A. et al. Optimizing the clinical application of rheumatology guidelines using large language models: A retrieval-Augmented Generation framework integrating EULAR and ACR recommendations. medRxiv (2025) doi:10.1101/2025.04.10.25325588.

25. Evangelista, E., Ruba, F., Bukhari, S. M. S., Nazir, A. & Sharma, R. S. Developing a GraphRAG-enabled local-LLM for Gestational Diabetes Mellitus. medRxiv (2025) doi:10.1101/2025.04.28.25326568.

26. Chronic kidney disease: assessment and management. NICE https://www.nice.org.uk/guidance/ng203.

2 A modular graph-based Retrieval-Augmented Generation (RAG) system. https://github.com/microsoft/graphrag.

28. Bastian, M., Heymann, S. & Jacomy, M. Gephi: An open source software for exploring and manipulating networks. Proceedings of the International AAAI Conference on Web and Social Media 3, 361–362 (2009).

29. Hu, Y. Efficient and High Quality Force-Directed Graph Drawing. Math. J. 10(1), 37-71., (2005).

30. Vincent D Blondel, Jean-Loup Guillaume, Renaud Lambiotte and Etienne Lefebvre. Fast unfolding of communities in large networks. Journal of Statistical Mechanics: Theory and Experiment.

31. Amugongo, L. M., Mascheroni, P., Brooks, S., Doering, S. & Seidel, J. Retrieval augmented generation for large language models in healthcare: A systematic review. PLOS Digit. Health 4, e0000877 (2025).

32. Miao, J., Thongprayoon, C., Suppadungsuk, S., Garcia Valencia, O. A. & Cheungpasitporn, W. Integrating retrieval-augmented generation with large language models in nephrology: Advancing practical applications. Medicina (Kaunas) 60, 445 (2024).

